# Label-free cell detection of acute leukemia using ghost cytometry

**DOI:** 10.1101/2023.07.14.23292646

**Authors:** Yoko Kawamura, Kayoko Nakanishi, Yuri Murata, Kazuki Teranishi, Ryusuke Miyazaki, Keisuke Toda, Toru Imai, Yasuhiro Kajiwara, Keiji Nakagawa, Hidemasa Matsuo, Souichi Adachi, Sadao Ota, Hidefumi Hiramatsu

## Abstract

Early diagnosis and prompt initiation of appropriate treatment are critical for improving the prognosis of acute leukemia. Currently, acute leukemia is diagnosed by microscopic morphological examination of bone marrow smears and flow cytometric immunophenotyping of bone marrow cells stained with fluorophore-conjugated antibodies. However, these diagnostic processes require trained professionals and are time and resource-intensive. Here, we present a novel diagnostic approach using ghost cytometry, a recently developed high-content flow cytometric approach, which enables machine vision-based, stain-free, high-speed analysis of cells, leveraging their detailed morphological information. We demonstrate that ghost cytometry can detect leukemic cells from the bone marrow cells of patients diagnosed with acute lymphoblastic leukemia (ALL) and acute myeloid leukemia (AML), without relying on biological staining. The method presented here holds promise as a precise, simple, swift, and cost-effective method for the diagnosis of acute leukemia in clinical practice.

## 1 INTRODUCTION

Acute leukemia is one of the most common types of cancer and remains a leading cause of death worldwide [1-3]. Early diagnosis is critically important to increase survival chances and improve prognosis. Acute leukemia is currently diagnosed by morphological examination of bone marrow (BM) smears using microscopy [4-5]. Immunophenotyping using flow cytometry (FCM) has recently been used for the diagnosis of acute leukemia in combination with the microscopic examination [5-10]. However, manual reading under microscopy requires skilled specialists and is often labor-intensive and time-consuming. While FCM allows a quantitative and high-speed analysis of cells, it relies on cell staining with fluorophore-conjugated antibodies, which may lead to errors due to a lack of cell morphological correlation, reduced antigen expression, and nonspecific binding. In addition, an FCM test with antibodies is expensive and its procedures and interpretations are often complicated, therefore limiting facilities that can perform it. Thus, a simple, accurate, and cost-effective method for the diagnosis of acute leukemia is desired.

Ghost cytometry (GC) is a recently developed high-content flow cytometric approach, which enables a machine vision-based, high throughput analysis of cells based on their detailed structural information [11, 12]. In a previous study, we have demonstrated that its label-free mode (label-free GC: LF-GC) can distinguish different cell states, cell types derived from human induced pluripotent stem (iPS) cells, and subtypes of peripheral white blood cells, without the use of biological cell staining [13]. However, its application to BM cells and its utility for diagnosis in clinical practice have not been investigated yet. This study demonstrates that LF-GC can detect leukemic cells in BM cells from patients diagnosed with acute lymphoblastic leukemia (ALL) and acute myeloid leukemia (AML), presenting that this approach is a promising candidate as a new diagnostic method for acute leukemia.

## 2 MATERIALS and METHODS

### 2.1 Cell line

The NB-4 cell line (human, B-cell leukemia cell line) was purchased from the German Collection of Microorganisms and Cell Cultures (DSMZ, Braunschweig, Germany). The cell line was cultured in the Roswell Park Memorial Institute (FUJIFILM Wako Pure Chemical, 189-02025) 1640 medium with 10% fetal bovine serum (Merck, F7524) and 1% penicillin/streptomycin (FUJIFILM Wako Pure Chemical, 168-23191) at 5% CO^2^ and 37°C.

### 2.2 ALL and AML patients

BM aspirates of two patients diagnosed with acute leukemia were collected. Patient 1 was a male diagnosed with B-cell acute lymphocytic leukemia. The May-Grunwald-Giemsa (MGG) stained-BM smear showed 60.0% blasts. The blast cells were small to medium-sized and had scanty basophilic cytoplasm with homogeneous fine chromatin. Patient 2 was a female diagnosed with overt acute myeloid leukemia and had a transplant for myelodysplastic syndromes (MDS). The MGG stained-BM smear showed 31.2% blasts. The blast cells were medium to slightly large-sized with a high nuclear/cytoplasm ratio with basophilic cytoplasm. They had soft fine chromatin with the nucleolus.

### 2.3 Sample preparations

NB-4 cells and a BM aspirate of a healthy donor were first fixed with a Lysing solution of 10× concentrate (BD bioscience, 349202): the cells were incubated with the 10-fold Lysing solution diluted with deionized water for 10 min. After fixation, the NB-4 cells were labeled with Fixable Far Red Dead Cell Stain (FFR, Thermo Fisher Scientific, L34974): the solid product of FFR was dissolved in dimethyl sulfoxide (DMSO, FUJIFILM Wako Pure Chemical, 046-21981) and the cells were incubated using the FFR DMSO stock solution 50-fold diluted with phosphate-buffered saline (PBS, FUJIFILM Wako Pure Chemical, 045-29795) solution containing 1% of Tween 20 (Merck, P1379-25ML) for 60 min. The BM cells were incubated with a DMSO solution diluted 50-fold with PBS containing 1% of Tween 20 (Merck, P1379-25ML) for 60 min so that the exposure to DMSO became equivalent between the NB-4 and BM cells. After staining, the NB-4 and BM cells were mixed at an approximately equal ratio and immediately introduced into a glass flow cell on a GC setup.

The BM aspirates of patients with ALL and AML were first lysed using the VersaLyse Lysing Solution (Beckman Coulter, A09777) for 15 min. The cells were then stained with the APC/Cyanine7 anti-human CD45 Antibody (BioLegend, 304014). The antibody was diluted 40-fold with PBS (FUJIFILM Wako Pure Chemical, 045-29795) solution containing 0.5% of Bovine Serum Albumin (Merck, A7906-100G) before use. After staining, the cells were fixed with Lysing solution 10× concentrate (BD bioscience, 349202) in the same manner as described above. After fixation, the cells were immediately introduced into a glass flow cell on a GC setup.

### 2.4 Experimental setup

Cells flowed through a glass flow cell (Hamamatsu Photonics Inc.) with a channel cross-section dimension of 150×150 µm^2^ at the measurement position. The sample flow was hydrodynamically focused into a narrow stream using sheath flows. The sheath fluid (IsoFlow, Beckman Coulter) was driven at a pressure of about 305 kPa using a homemade pressure pump and the sample fluid was driven at a flow rate of approximately 10 to 20 µl/min using a syringe pump (KD scientific). The GC set-up equips three colors of continuous-wave lasers: a 405-nm-wavelength violet laser, a 488-nm-wavelength blue laser, and a 637-nm-wavelength red laser. The violet laser which passes through a diffractive optical element forms a structured illumination pattern on the focal plane of an objective lens. The blue and red laser beams are focused using objective lenses in spots as in a conventional flow cytometer.

When cells flow through a structured light illumination in a microfluidic channel, compressed imaging signals are acquired as temporal waveforms encoding morphological information of individual cells, which we call ghost motion imaging (GMI) waveforms [11-13] (Figure 1A). In this work, we utilized two label-free GMI (LF-GMI) modalities called diffractive GMI (dGMI) waveform and forward-scattering GMI (fsGMI) waveform, which are analogous to diffractive and forward-scattering darkfield microscopy images, respectively. The dGMI and fsGMI waveforms, as well as conventional flow cytometric measures such as forward scattering (FSC), backscattering (BSC), and red fluorescence signals at a 637 nm wavelength were acquired by photomultiplier tubes (PMT, Hamamatsu Photonics Inc.). The field-programmable gate array development board (FPGA, TR4, Terasic) with a homemade analog/digital converter recorded a fixed length of signal segments simultaneously from each PMT channel with a fixed trigger condition applied to the FSC signal.

**FIGURE 1:**
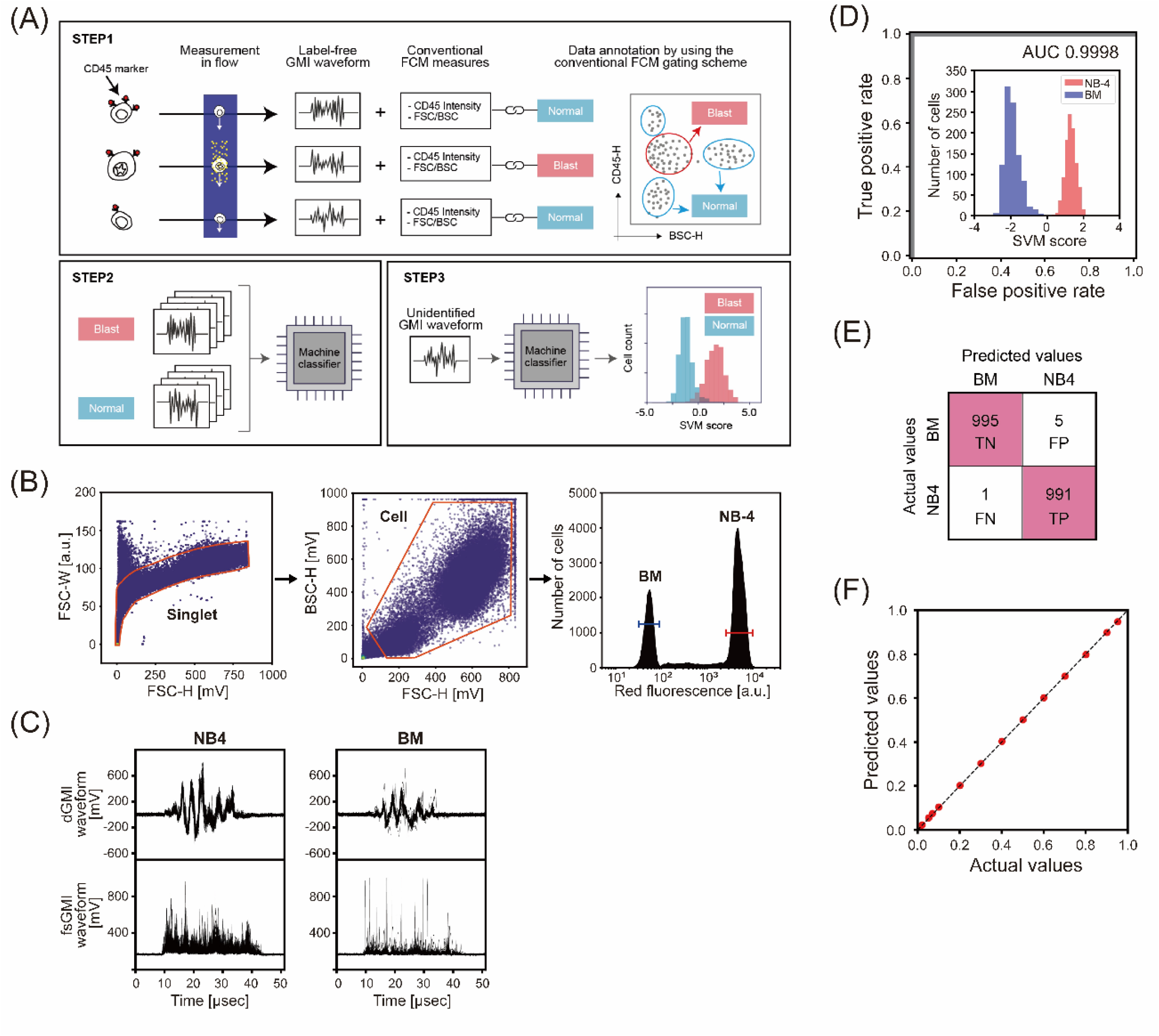
Label-free detection of NB-4 cells spiked in BM cells from a healthy donor. **(A)** A workflow for the label-free detection of blast cells in BM using LF-GC. STEP1, LF-GMI waveforms as well as the conventional flow cytometric measures such as forward-scattering (FSC), back-scattering (BSC), and fluorescence signals as ground truth labels are collected using GC. Homemade software is used for data collection. Subsequently, the LF-GMI waveforms of blast cells and normal cells in BM are identified (annotated) using ground truth labels defined by the widely used CD45 gating schemes in the detection of acute leukemia by FCM (upper panel). STEP2, a classifier based on a machine-learning algorithm, the support vector machine (SVM) in this study, is trained using the annotated LF-GMI waveforms (lower left panel). STEP3, the classifier predicts the class (blast or normal) of cells based solely on unidentified LF-GMI waveforms, a test dataset that was not used to develop the classifier, without using biological labels. **(B-F)** Detection of NB-4 cells spiked in the BM of a healthy donor. **(B)** Gating strategies applied to identify (annotate) NB-4 and BM cells for training a classifier. Singlets were selected in the FSC-H (height)/FSC-W (width) plots (left panel). Debris was excluded and cells were selected in the FSC-H/BSC-H plots (center panel). NB-4 and BM cells were identified (annotated) based on the intensities of red fluorescence stained as a ground truth label (right panel). **(C)** Representative LF-GMI waveforms, pairs of dGMI and fsGMI waveforms, corresponding to NB-4 and BM cells after annotation. Fifty waveforms were overlapped in each panel. For training a classifier, 6,000 pairs of LF-GMI waveforms (3,000 pairs for each class) were used. **(D)** An ROC curve and SVM score histogram (the inset figure) in classifier evaluation; 2,000 pairs of LF-GMI waveforms (1,000 pairs for each class) were used for validation. The AUC score of 0.9998 was obtained. Red and blue colors at the inset of the figure correspond to NB-4 and BM cells defined by the red fluorescence intensities shown in Figure 1B, respectively. **(E)** A confusion matrix in classifier evaluation when the decision boundary of SVM scores was 0 (TP, true positive; FP, false positive; TN, true negative; FN, false negative). **(F)** Detection of NB-4 cells spiked in BM cells at the concentration ratios of 1–95%. Data from NB-4 cells and BM cells were computationally mixed and analyzed. 5,000 pairs of LF-GMI waveforms were used in each condition. The decision boundary of SVM scores was 0. The dashed line shows x = y.

### 2.5 Data analysis using machine learning

For high-speed and accurate information processing, LF-GC directly analyzes the compressed LF-GMI (dGMI and fsGMI) waveforms using a machine-learning algorithm without the need for computationally intensive digital image production. Conventional flow cytometric data modalities such as FSC, BSC, and red fluorescence signals from biological labeling were utilized for annotation in supervised machine-learning analysis. Specifically, the biological labels were used as ground truth labels to identify (annotate) blast and normal cells in BM based on flow cytometric gating schemes. The annotated LF-GMI waveforms were subsequently used for training a classifier based on the support vector machine (SVM) algorithm. The classification abilities of the trained models were evaluated by comparing predicted results with the ground truth labels based on a receiver operating characteristic (ROC) curve, the area under the ROC curve (AUC), and the confusion matrix. Finally, by applying the trained classifier to unidentified LF-GMI waveforms, a test dataset that was not used to develop the classifier, we demonstrated the detection of blast cells in BM without using biological labels.

## 3 RESULTS and DISCUSSION

### 3.1 Detection of NB-4 cells in BM cells from a healthy donor

We first detected NB-4 cells experimentally spiked in the BM cells of a healthy donor including immature hematopoietic cells that appear morphologically similar to leukemic cells (Figure 1B-F). Before the measurement, we stained only NB-4 cells with a red fluorescence marker as a ground truth label as described above. After data collection, we identified (annotated) NB-4 and BM cell populations through gating based on the red fluorescence intensities (Figure 1B). Representative LF-GMI waveforms obtained for NB-4 and BM cells after annotation are shown in Figure 1C. Using the LF-GMI waveforms (annotated), we then trained a classifier and evaluated its performance. Figure 1D shows an ROC curve with an AUC score of 0.9998 in classifier evaluation, exhibiting a high classification ability. Figure 1E depicts a confusion matrix when the decision boundary of SVM scores was 0 (SVM score ≥ 0, predicted as blast; SVM score < 0, predicted as normal), resulting in a high accuracy value of 0.997. Furthermore, to test the robustness of the model, we detected NB-4 cells spiked in BM cells at various concentration ratios (1–95%) using the classifier. As shown in Figure 1F, the LF-GC-based detection of the acute leukemia cell line consistently showed high performance over a wide range of concentration ratios.

### 3.2 Detection of blast cells in BM cells from patients with acute leukemia

Next, we detected blast cells in the BM of patients diagnosed with acute leukemia. BM aspirates were collected from two patients diagnosed with B-cell ALL and AML, as described above. Before LF-GC measurements, we stained the BM samples with a CD45 surface marker as a ground truth label. After data collection, we identified (annotated) blast and normal cells in the BM samples according to the commonly used CD45 gating strategies [14,15] (Figure 2A). Representative LF-GMI waveforms obtained for NB-4 and BM cells after annotation are shown in Figure 2B. Subsequently, we trained and evaluated classifiers using the annotated LF-GMI waveforms. Figure 2C shows ROC curves in classifier evaluations, showing high AUC scores of 0.986 for ALL and 0.941 for AML. Figure 2D depicts confusion matrices in classifier evaluations when the decision boundary of SVM scores was 0 (SVM score ≥ 0, predicted as blast; SVM score < 0, predicted as normal), showing high accuracy values of 0.948 and 0.878 for ALL and AML, respectively.

**FIGURE 2:**
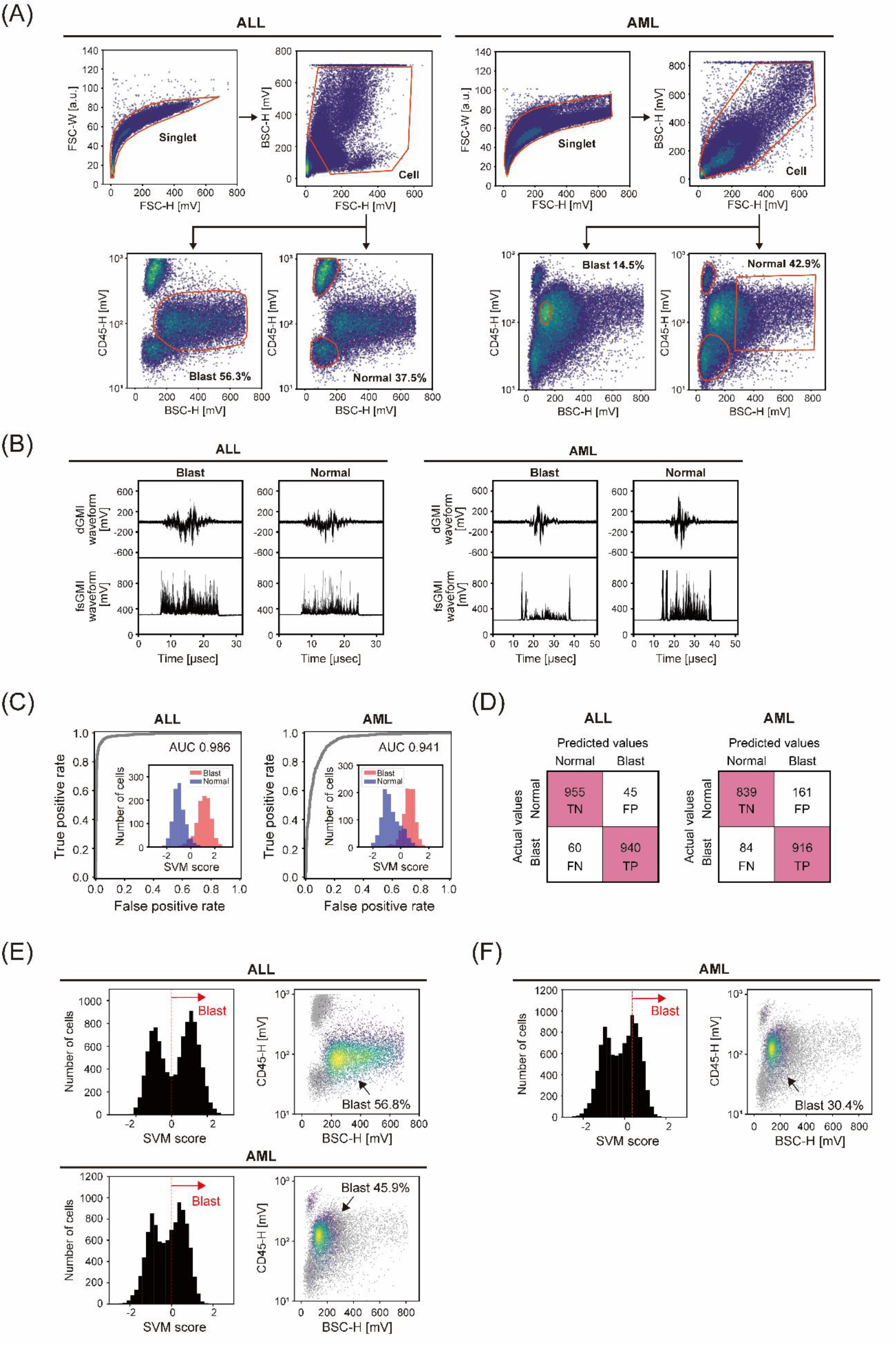
Label-free detection of blast cells in BM cells from patients diagnosed with ALL and AML. **(A)** Gating strategies applied to identify (annotate) blast cells and normal cells in BM for training a classifier. Singlets were selected in the FSC-H/FSC-W plots (upper left panels). Debris was excluded and cells were selected in the FSC-H/BSC-H plots (upper right panels). Blast and normal cells were identified (annotated) in the CD45/BSC plots based on the conventional CD45 gating schemes (lower panels). **(B)** Representative LF-GMI waveforms, pairs of dGMI and fsGMI waveforms, corresponding to blast cells and normal cells in the BM samples after annotation. Fifty waveforms were overlapped in each panel. In each ALL and AML case, 6,000 pairs of dGMI and fsGMI waveforms (3,000 pairs for each class) were used for training a classifier. **(C)** ROC curves and SVM score histograms (the inset figures) in classifier evaluations. Red and blue colors in the inset figures correspond to blast and normal cells defined by gating shown in Figure 2A, respectively. In each case, 2,000 pairs of LF-GMI waveforms (1,000 pairs for each class) were used for the validation. The AUC scores of 0.986 and 0.941 were obtained for ALL and AML, respectively. **(D)** Confusion matrices in classifier evaluations when the decision boundary of SVM scores was 0. **(E-F)** Histograms and BSC/CD45 back plots of the SVM scores obtained by applying the classifiers to unidentified (non-annotated) LF-GMI waveforms; 10,000 pairs of LF-GMI waveforms were classified in each case. In **(E)**, the decision boundary of SVM scores was 0 in both cases. In **(F)**, the decision boundary of SVM scores was 0.37 only in the AML case. Prediction as blast and normal cells are depicted as color density and light gray plots, respectively, in the right panels.

Finally, we applied the developed classifiers to unidentified (non-annotated) LF-GMI waveforms, a test dataset that was not used to develop the classifier (Figures 2E,F). The SVM score histograms of the test datasets (Figures 2E,F, left panels) exhibit bimodal peaks corresponding to blast and normal cells for both cases. Furthermore, there is a good agreement between the blast regions defined by LF-GC-based predictions and those defined by the CD45 gating methods (Figures 2E,F, right panels). When the decision boundary of SVM scores was set to 0, the blast ratios were calculated to be 56.8% for ALL and 45.9% for AML (Figure 2E). Based on the May-Grunwald-Giemsa staining, morphological examinations of the same cases by medical technologists at diagnosis determined the blast ratios to be 60.0% for ALL and 31.2% for AML. In the case of ALL, the blast ratio obtained by LF-GC is thus highly consistent with the result of the manual diagnosis. In the case of AML, the blast ratio obtained by LF-GC was higher than that by manual examination. We think that the observed result is reasonable as we excluded cell populations with intermediate morphological characteristics between blast and normal cells from training data sets. Given the heterogeneity of AML cells, we gated 14.5% of cells as blast cells and 42.9% as normal cells and excluded the remaining 42.6% from the gating scheme (Figure 2A, right bottom panels). Such a tight-gating approach allows us to build a model that learns the distinct morphological characteristics of each class for non-intermediate morphology cells. On the other hand, this model was trained to maximize the AUC score when a decision boundary set at an SVM score was 0. This approach does not fully reflect the criteria employed by human examiners when classifying the morphologically heterogeneous intermediate cells under microscopy. To fill this gap, we introduced a new decision boundary set at an SVM score of 0.37 (Figure 2F, left panel). Under this revised criterion, cells with an SVM score of 0.37 or higher are identified as blast cells, while cells with a score below this threshold are identified as normal cells. Redefining the criteria effectively narrows down the cells more likely to be blast cells from the entire cell population, including intermediate cells. The results of this optimization are reflected in the CD45/BSC back plot, which shows a more refined and narrow blasts region (Figure 2F, right panel). As a result, the blast ratio is reduced to 30.4%, a figure that closely matches the results obtained from human-based testing. This result indicates the possibility of quantitatively aligning the degree of morphological abnormality judged by LF-GC with that by humans, even for AML cells with morphological heterogeneity. We believe that the accumulation of additional leukemia cases and cell data in combination with metadata such as detailed clinical and demographic patient background information will lead to the development of a robust, accurate, and reliable protocol for the application of LF-GC in the diagnosis of acute leukemia. The machine vision-based method presented here is expected to be applied as an accurate, simple, rapid, and economical diagnostic method of acute leukemia in a clinical setting.

## Data Availability

The data that support the findings of this study are available from the corresponding author upon reasonable request.

## ACKNOWLEDGEMENTS

We thank all members of ThinkCyte K.K., in particular, Keiki Sugimoto, Hiroaki Adachi, and Hikari Morita, for their discussions. We thank the patients and their families.

## AUTHOR CONTRIBUTIONS

Y.Kawamura, K.Nakanishi, H.M., S.A., S.O., and H.H. conceptualized the study; K.Nakanishi provided the cell line and patient samples; Y.Kawamura and R.M. performed cell assays; K.Toda and T.I. developed the GC optical unit; Y.Kajiwara developed the GC software; K.Nakagawa developed the fluidic unit and integrated the GC set-up; Y.Kawamura conceived, designed, and performed the GC experiments, data analysis, and data curation; Y.M. and K.Teranishi performed GC experiments and data analysis; S.O. and H.H. supervised the work; Y.Kawamura, K.Nakanishi, H.M., S.A., S.O., and H.H. discussed the results and interpreted the data; Y.Kawamura, S.O., K.Nakanishi, and H.H. wrote the manuscript with contributions from all authors.

## CONFLICT OF INTEREST STATEMENT

S.O. is the founder and shareholder of ThinkCyte K.K., a company engaged in developing GC-based cell sorting; Y.Kawamura, Y.M., K.Teranishi, R.M., K.Toda, T.I., and K.Nakagawa have shares of stock options of ThinkCyte K.K.; S.O., Y.Kawamura, K.Toda, T.I., and K.Nakagawa have filed patent applications related to this paper.

## Notes

### Funding Statement

This study did not receive any funding.

### Author Declarations

Kyoto University Graduate School and Faculty of Medicine, Ethics Committee gave ethical approval for this work.

